# Baseline TyG index, myoglobin, and cerebral infarction history predict the onset of pulmonary hypertension in coronary artery disease patients after PCI treatment within a median of 4.5 years: a prospective cohort study

**DOI:** 10.1101/2023.02.21.23286276

**Authors:** Li Xie, Shilin Fu, Yuzheng Xu, Litong Ran, Jing Luo, Rongsheng Rao, Jianfei Chen, Shi-Zhu Bian, Dehui Qian

## Abstract

**Aim:** To identify the predictive role of the TyG index for the onset of pulmonary hypertension in patients with coronary artery disease (CAD) who underwent percutaneous coronary intervention (PCI) treatment.

**Methods:** We performed this prospective cohort study among CAD patients who received PCI treatment in our center from July 2016 to October 2022. The baselines of echocardiography at both cross-sections and blood biomarkers. A coronary angiography operation was also performed. Within a median of 4.5 years of follow-up, the patients underwent echocardiography to measure their pulmonary hypertension (PH).

**Results:** Baseline BNP was statistically higher in the PH patients (p = 0.007). The baseline myoglobin (MYO), was significantly higher among PH patients (p < 0.001). Though the glucose level showed no difference between PH and non-PH groups, the HDL-C was in a lower level in the PH group (p = 0.033). However, TyG index showed no differences between PH and non-PH groups [6.95 (6.47-7.36) vs. 7.15 (6.49-7.96), p = 0.202]. In the univariate regression, cerebral infarction history, right atria end-diastolic internal diameter, MYO, triglyceride, HDL-C and TyG index (p < 0.05) were potential predictors for PH. Finally, the adjusted logistic regression indicated that cerebral infarction history (p = 0.39), MYO (p = 0.044) and TyG index (p = 0.048) were independent predictors of the onset of PH.

**Conclusion:** PH is prevalent in CAD patients after PCI treatment. The baseline TyG index, cerebral infarction history, and MYO level were independent predictors for PH in CAD patients after PCI treatment.

## 1 Introduction

Insulin resistance (IR) has been considered a critical pathophysiological process in cardiometabolic diseases and other cardiovascular diseases in various clinical and basic research (1,2). The triglyceride-glucose (TyG) index is the product of fasting plasma glucose (FPG) and triglycerides (TGs). It has been indicated to be a reliable surrogate marker of IR (3,4). Furthermore, it has been indicated that the TyG index is well-correlated to the homeostasis model which is used to assess IR (2,5,6). It has also been used as a marker and predictor for a variety of diseases, including major adverse cardiovascular events (MACE) as well as atrial fibrillation (AF), over traditional risk factors (5,7-9). The TyG index has been used widely because IR is a leading reason for the elevations of triglyceride and glucose among healthy populations.

Among ST elevation myocardial infarction patients, the higher TyG index has been indicated to increase the risk of major adverse cardiac and cerebrovascular events(7,10,11). Although a number of the previous studies have indicated the associations between TyG index and vascular diseases, no studies further identify the predictive role of the TyG index in pulmonary hypertension (PH) among coronary artery disease (CAD) patients after PCI treatment (9,12-14).

The complications of CAD patients after PCI such as MACE and heart failure have received sufficient attention (15-18). However, the effect of CAD on pulmonary circulation has not been investigated widely, especially after patients received PCI treatment (19,20).

PH has been considered as one of the most common complications in patients with cardiovascular diseases. PH is characterized with an elevated pulmonary arterial pressure (PAP) and significant remodeling of pulmonary arterioles (21-23). PH is a clinical syndrome which is caused by various etiologies resulting in modifications of function or structures of pulmonary vasculature, leading to the rise of PAP and pulmonary vascular resistance (PVR) (21). It has been indicated that PH is the primary reason for right heart failure, which may be a fatal complication in patients with cardiovascular disease. The incidence of PH in various cardiovascular diseases has been reported to be from 20 to 80%. Except in cases of congenital heart disease and dilated cardiomyopathy, PH was frequently found in CAD patients (19). Recently, PH has also been recognized as a threat in cardiovascular patients which may lead to fatigue, shortness of breath and even right heart failure and death. Because the onset of PH among patients is often obscured by other diseases, when PH is diagnosed, it may be in its later stage and difficult to treat. Thus, the early discovery and diagnosis of PH is of great importance (24).

It is well known that PH is a multi-factorial and complex clinical syndrome which accompanies a variety of diseases including heart failure, lung diseases, hypoxia and chronic thromboembolism (22). PH in CAD patients after PCI has not been identified (25). We designed this study to identify the predictive role of the TyG index in PH populations who suffers from CAD patients after PCI operation treatments.

## 2 Methods

### 2.1 Patients and Study Design

All consecutive patients who were clinically diagnosed with CAD in our center from July 2016 and July 2019 were included into this study according to the inclusion and exclusion criteria. The follow-up was finished in October 2022. The inclusion criteria were: aged from 18-80 years, and clinically diagnosed with unstable angina. A patient with one of the following diseases including aortic aneurysm, chronic kidney diseases, infections, congenital heart diseases, cardiac valve diseases, connective tissue diseases, and cancer. A patient who has received PCI or coronary artery bypass graft previously has also been excluded.

Finally, a total of 247 patients with CAD were enrolled. Upon admission, the patients’ laboratory and demographic data as well as the disease’ history have been tested and recorded. Our study conforms to the Declaration of Helsinki. The study has been approved by our ethical committee of Xinqiao Hospital, Third Military Medical University (Army Military Medical University). The patients were familiarized with the procedure and contents of the study and provided written informed consent.

Venous blood samples were obtained from patients on admission prior to coronary intervention. Both serum and plasma samples were obtained and placed on ice. After they were centrifuged within 30 min, they were stored at −70 °C for the further test.

### 2.2 The Assessments of CAD Severity for each patient

We have performed a scheduled coronary angiography (CAG) for each patient according to standard techniques. At least two of our trained interventional cardiologists. CAD in our center was defined as following: at least a single-vessel with a stenosis ≥50%.

### 2.3 Biochemical Parameters

We have obtained venous blood samples from CAD patients in the early morning after at least 12 hours’ overnight fast.

We have performed the biochemical examinations. We have tested the levels of blood glucose (fasting blood glucose, FBG) and lipids including total cholesterol (TC), high-density lipoprotein cholesterol (HDL-C), low-density lipoprotein cholesterol (LDL-C) and triglyceride (TG) concentrations by using Beckman Coulter AU5800 system (USA). Furthermore, we have also detected the liver and renal functions with the same machine above.

We have also measured the plasma UA with Roche enzymatic assays (Roche Diagnostics GmbH, Mannheim, Germany). Furthermore, plasma creatinine (Cr) concentration was also measured by using Hitachi 7600 autoanalyzer (Hitachi, Tokyo, Japan) via enzymatic assay. The blood urea nitrogen concentration (BUN) was also measured (FERENE methods, Beckman AU5821).

Finally, we have tested the electrolytes including sodium concentration (Na^+^), serum calcium concentration (Ca^2+^), phosphate concentration (P), serum potassium concentration (K^+^) and chloride ion (Cl-) were also measured.

The levels of myohemoglobin (MYO), creatine kinase isoenzyme-MB (CK-MB) and cardiac troponin I (TnI) in serum were measured using standard assay kits and a Geteln 110 (Jiangsu, China) automatic chemiluminescence immunoassay analyzer, and the plasma level of brain natriuretic peptide (BNP) was also measured according to the manufacturer’s instructions using a Geteln 110 (Jiangsu, China) automatic chemiluminescence immunoassay analyzer.

### 2.4 Clinical data collection

The patients were followed-up by our physicians. We used a standardized self-report questionnaire form to record patients’ data including demographic information and diseases history as well as the medication use. Height and weight were also measured. The history of smoking status and drinking.

The systolic blood pressure (SBP), heart rate (HR) and diastolic blood pressure (DBP) were measured on the right arm in a sitting position after sufficient rest.

### 2.5 Echocardiography examinations

Both at the baseline and follow-up cross-sections, echocardiography examinations have been performed by our trained technicians with an ultrasonography workstation (CX50, Philips, USA; probe: S5). The patients were positioned in a supine position after a rest of ten min. The technicians have measured and recorded the end-diastolic internal diameters of the left and right atria (LAD and RAD). As regulation, the end-diastolic internal diameters of left and right ventricles (LVDD and RVD) have also been measured and recorded. In the next step, the thickness of the interventricular septum (IVS) and left ventricular posterior wall (LVPW) have also been evaluated and recorded. The fractional shortening (FS), stroke volume (SV) and ejection fraction (LVEF) has been measured and calculated. Finally, the tricuspid regurgitation area (TRA), TR velocity (TRV) which was used to calculate the PAP, and TR pressure.

### 2.6 Variables

We measured and calculated the systolic PAP (sPAP) by using the modified Bernoulli equation using the tricuspid systolic jet according to the American Society of Echocardiography:

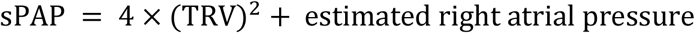

The estimated right atrial pressure was recorded as 5 mmHg (normal right atrium size), 10 mmHg (mildly enlarged right atrium size), and 15 mmHg (significantly enlarged right atrium size) in according to the right atrium size.

An sPAP >35 mmHg was defined as PH. In contrast, the patients with sPAP <35 mmHg were included in the non-PH group According to the American Society of Echocardiography and other studies.

### 2.7 Follow-up

The follow-up at the outpatient department was performed by our doctors, including Li Xie, Shi-Zhu Bian, Dehui Qian, Shilin Fu and Jianfei Chen, with an objective case report form, and regular examinations for the CAD patients. Furthermore, the examination of echocardiograms was performed by a technician, Dr. Rongsheng Rao.

### 2.8 Statistical Analysis

All of our statistical analyses were employed with SPSS software (version 26.0, IBM Corp., America). Continuous normalized distribution variables are expressed as mean ± standard deviation (SD) and were compared using Students’ test between two groups. The non-normalized distributed data are summarized as medians (interquartile ranges). Each categorical variable was expressed in case and proportions. The differences of the non-normalized distributed data between the PH group and non-PH group were analyze by using a Kruskal-Wallis nonparametric test. The correlations between baseline parameters and sPAP were identified by using Spearman’s correlation analyses.

The potential predictors were analyzed by univariate and multivariate regressions. The backward stepwise logistic regression models were used to identify the predictors for the onset of PH in CAD patients. Statistical significance was defined as a p value is less than 0.05.

## 3 Results

Within a median of 4.5 years’ follow-up, the incidence of PH was 8.91%. The demographic data were shown in table 1. The LAD and LVDD increased dramatically while the LVEF decreased significantly (p < 0.001). Furthermore, the right heart structure also changed significantly (table 2). However, at the baseline cross-section, no significant differences were found between the PH and non-PH groups among LAD, LVDD, RAD, RVD, LVEF and SV (all p values are greater than 0.05). The biomarker of heart failure, BNP, was dramatically higher in the PH population than in the non-PH ones (68.00 [18.05-196.00] vs. 174.00 [76.25-447.75], p = 0.007). The biomarkers for cardiac injury, including TNI and CK-MB, showed no significant differences between PH and non-PH groups (p > 0.05). However, baseline MYO was dramatically higher in the PH group than that in the non-PH populations (56.30 [42.65-77.25] vs.76.30 [64.38-126.35] ng/ml, p < 0.001). We also found that HDL-C (1.06 [0.870-1.26] vs. 0.915 [0.770-1.071] mmol/L) was significantly lower in the PH group than that in the non-PH group (p = 0.033). However, the TyG index showed no significant difference between PH patients and non-PH ones (6.95 [6.47-7.36] vs. 7.15 [6.49-7.96], p = 0.202). The basic parameters, including HR, SBP and DBP, showed similar levels between PH group and non-PH group (p > 0.05) (table2-3).

**Table 1.** The characteristics of the patients

|  | Overall<br>(n = 247) | Non-PH<br>(n = 225) | PH<br>(n = 22) | p value |
| --- | --- | --- | --- | --- |
| Age (years) | 68.84±12.12 | 68.54±12.16 | 71.91±11.51 | 0.214 |
| BMI (kg/m<sup>2</sup>) | 24.10±3.42 | 24.18±3.49 | 23.22± | 0.206 |
| Gender, female, n (%) | 92(37.2%) | 85(37.8%) | 7(31.8%) | 0.581 |
| Smoking, n (%) | 104(42.1%) | 97(43.1%) | 7(31.8%) | 0.306 |
| Drinking n (%) | 43(17.4%) | 40(17.8%) | 3(13.6%) | 0.775 |
| Cerebral infarction history n (%) | 55(22.3%) | 46(20.4%) | 9(40.9%) | 0.028 |
| Hypertension n (%) | 158(64%) | 144(64.0%) | 14(63.6%) | 0.490 |
| T2DM, n (%) | 70(28.3%) | 64(28.4%) | 6(27.3%) | 0.927 |
| Meditation |  |  |  |  |
| CCB, n (%) | 50(20.2%) | 45(20.0%) | 5(22.7%) | 0.573 |
| ACEI/ARB, n (%) | 175(70.9%) | 161(71.6%) | 16(72.7%) | 0.475 |
| β-blocker, n (%) | 175(70.9%) | 161(71.6%) | 14(63.6%) | 0.884 |
| Diuretic, n (%) | 55(22.3%) | 50(22.2%) | 5(22.7%) | 0.784 |

The Cerebral infarction history showed significant difference between PH and non-PH groups.
ACEI: angiotension converting enzyme inhibitor; ARB: angiotensin receptor blocker; CCB: calcium channel blockers;T2DM: diabetes mellitus type 2.

**Table 2.** Hemodynamics changes from baseline to follow-up

|  | Baseline | Follow up | p |
| --- | --- | --- | --- |
| LAD (mm) | 35.69±5.00 | 35.74±4.59 | <0.001 |
| LVDD (mm) | 47.02±6.02 | 47.73±6.08 | <0.001 |
| LVEF (%) | 61.12±9.91 | 61.03±8.68 | <0.001 |
| SV (ml) | 70.32±18.59 | 70.35±20.06 | 0.241 |
| RAD (mm) | 35.45±4.28 | 35.38±3.64 | <0.001 |
| RVD (mm) | 34.02±3.21 | 33.79±3.82 | <0.001 |
| TRA (cm<sup>2</sup>) | 0.00(0.00-1.00) | 0.50(0.00-1.00) | 0.127 |
| TRV (m/s) | 0.00(0.00-243.00) | 200.00(0.00-240.00) | 0.573 |

The parameters of hemodynamics changed significantly from baseline to the follow-up.

**Table 3.** Differences between PH and non-PH patients

|  | Non-PH patients<br>(n = 225) | PH patients<br>(n = 22) | P value |
| --- | --- | --- | --- |
| Echocardiographic parameters |  |  |  |
| LAD (mm) | 35.52±4.97 | 37.36±5.09 | 0.100 |
| LVDD (mm) | 46.89±5.71 | 48.31±8.64 | 0.290 |
| LVEF (%) | 61.29±9.57 | 59.41±13.084 | 0.396 |
| SV (ml) | 70.04±18.57 | 73.13±18.91 | 0.457 |
| RAD (mm) | 35.33±2.00 | 36.68±5.04 | 0.158 |
| RVD (mm) | 33.95±3.15 | 34.81±3.78 | 0.226 |
| FS (%) | 33.72±7.69 | 32.23±8.61 | 0.292 |
| TRA (cm<sup>2</sup>) | 0.00(0.00-1.00) | 0.500(0.00-1.52) | 0.512 |
| TRV (m/s) | 0.00(0.00-243.38) | 100.00(0.00-245.00) | 0.736 |
| Biomarkers |  |  |  |
| BNP (pg/ml) | 68.00 (18.05-196.00) | 174.00 (76.25-447.75) | 0.007 |
| TNI (ng/ml) | 0.05(0.05-0.05) | 0.05(0.05-0.05) | 0.307 |
| MYO (ng/ml) | 56.30 (42.65-77.25) | 76.30(64.38-126.35) | <0.001 |
| CK-MB (ng/ml) | 1.00(1.00-1.00) | 1.00(1.00-1.00) | 0.808 |
| Electrolytes |  |  |  |
| Mg<sup>2+</sup> (mmol/L) | 0.966±0.132 | 0.95±0.116 | 0.700 |
| K<sup>+</sup> (mmol/L) | 3.98±0.73 | 3.90±0.47 | 0.642 |
| Ca<sup>2+</sup> (mmol/L) | 2.200 (2.12-2.30) | 2.18 (2.11-2.26) | 0.705 |
| Na<sup>+</sup> (mmol/L) | 139.38±4.80 | 139.34±3.88 | 0.970 |
| Cl<sup>-</sup> (mmol/L) | 105.68±5.08 | 104.97±3.96 | 0.552 |
| TCO<sub>2</sub> (mmol/L) | 24.70±5.87 | 25.14±2.44 | 0.732 |
| P (mmol/L) | 1.11±0.49 | 1.08±0.30 | 0.843 |
| Renal function |  |  |  |
| UA(μmol/L) | 369.70 (304.55-442.30) | 355.05 (282.30-444.47) | 0.668 |
| Cr (μmol/L) | 78.30 (67.30-93.85) | 79.95 (68.45-93.90) | 0.818 |
| BUN (μmol/L) | 7.12±6.38 | 7.77±4.57 | 0.645 |
| eGFR(mL/min/1.73m<sup>2</sup>) | 81.12 (61.10-92.44) | 78.88 (60.25-85.46) | 0.445 |
| Serum lipids and glucose |  |  |  |
| TC (mmol/L) | 3.74(2.96-4.52) | 3.72 (2.97-4.19) | 0.597 |
| TG (mmol/L) | 1.25 (0.92-1.80) | 1.49 (0.35-2.42) | 0.541 |
| HDL-C (mmol/L) | 1.06 (0.870-1.26) | 0.915 (0.770-1.071) | 0.033 |
| LDL-C (mmol/L) | 2.31 (1.74-2.96) | 2.215 (1.780-2.59) | 0.433 |
| Glu (mmol/L) | 4.74 (4.20-5.52) | 5.32 (4.32-6.48) | 0.126 |
| TyG index | 6.95(6.47-7.36) | 7.15(6.49-7.96) | 0.202 |
| HbA1c (%) | 6.30(5.90-6.75) | 6.48 (6.075-7.00) | 0.220 |
| Basic parameters |  |  |  |
| HR (mmHg) | 72.54±14.50 | 76.20±22.01 | 0.285 |
| SBP (mmHg) | 127.55±19.24 | 125.92±17.63 | 0.702 |
| DBP (mmHg) | 72.67±13.06 | 72.03±12.91 | 0.828 |

HDL-C showed significant difference between PH and non-PH groups. However, the TyG index showed no significant differences between them.

In the next step, we analyzed the correlation between sPAP and all of the parameters. The demographic variables, including BMI and age, showed no close association with sPAP. Whilst we found that the baselines of TRA (r = 0.151, p = 0.017) and TRV (r = 0.156, p = 0.014) were significantly positively correlated with sPAP, other baseline hemodynamics parameters showed no close relationship with sPAP (table 4).

**Table 4.** Relationship between sPAP and other paramters

|  | Correlation coefficient r | P value |
| --- | --- | --- |
| *Demographic data* |  |  |
| Age (years) | 0.071 | 0.264 |
| BMI ((kg/m<sup>2</sup>)) | -0.009 | 0.884 |
| *Echocardiographic parameters* |  |  |
| LAD (mm) | 0.110 | 0.083 |
| LVDD (mm) | 0.060 | 0.347 |
| LVEF (%) | 0.071 | 0.268 |
| SV (ml) | 0.032 | 0.618 |
| RAD (mm) | 0.095 | 0.138 |
| RVD (mm) | 0.092 | 0.149 |
| FS (%) | 0.069 | 0.283 |
| TRA (cm<sup>2</sup>) | 0.151 | 0.017 |
| TRV (m/s) | 0.156 | 0.014 |
| *Biomarkers* |  |  |
| BNP (pg/ml) | 0.113 | 0.078 |
| TNI (ng/ml) | -0.041 | 0.519 |
| MYO (ng/ml) | 0.109 | 0.089 |
| CK-MB (ng/ml) | 0.040 | 0.531 |
| *Electrolytes* |  |  |
| Mg<sup>2+</sup> (mmol/L) | 0.034 | 0.490 |
| K<sup>+</sup> (mmol/L) | -0.006 | 0.924 |
| Ca<sup>2+</sup> (mmol/L) | 0.049 | 0.440 |
| Na<sup>+</sup> (mmol/L) | -0.059 | 0.352 |
| Cl<sup>-</sup> (mmol/L) | -0.074 | 0.247 |
| TCO<sub>2</sub> (mmol/L) | 0.061 | 0.338 |
| P (mmol/L) | -0.064 | 0.318 |
| *Renal function* |  |  |
| UA(μmol/L) | -0.003 | 0.968 |
| Cr (μmol/L) | -0.010 | 0.880 |
| BUN (μmol/L) | 0.029 | 0.650 |
| eGFR(mL/min/1.73m<sup>2</sup>) | 0.053 | 0.405 |
| *Serum lipids and glucose* |  |  |
| TC (mmol/L) | -0.089 | 0.166 |
| TG (mmol/L) | -0.105 | 0.099 |
| HDL-C (mmol/L) | -0.041 | 0.526 |
| LDL-C (mmol/L) | -0.117 | 0.066 |
| Glu (mmol/L) | -0.024 | 0.703 |
| TyG index | -0.095 | 0.136 |
| HbA1c (%) | -0.027 | 0.6720 |
| Basic parameters |  |  |
| HR (bpm) | -0.034 | 0.592 |
| SBP (mmHg) | -0.070 | 0.271 |
| DBP (mmHg) | -0.087 | 0.173 |

Baselines of TRA and TRV were closely correlated to the followed-up sPAP.

In the univariate regression, we performed analyses with each variable. We identified that cerebral infarction history (β = 0.991, odds ratio (OR): 2.694, 95% confidence interval (CI): 1.085-6.690, p = 0.033), LAD (β = 0.074, OR: 1.077, 95% CI: 0.987-1.174, p = 0.095), RAD (β = 0.055, OR: 1.057, 95% CI: 0.976-1.144, p =), MYO (β = 0.007, OR: 1.007, 95% CI: 1.001-1.013, p = 0.024), TG (β = 0.304, OR: 1.355, 95% CI: 1.062-1.730, p = 0.015), HDL-C (β = -1.744, OR: 0.175, 95% CI: 0.027-1.115, p = 0.065), HbA1c (β = 0.288, OR: 1.334, 95% CI: 0.975-1.825, p = 0.071) and TyG index (β = 0.628, OR: 2.389, 95% CI: 1.308-4.363, p = 0.005) were potential predictors/risk factors for PH in CAD patients after PCI treatment (table 5).

**Table 5.**
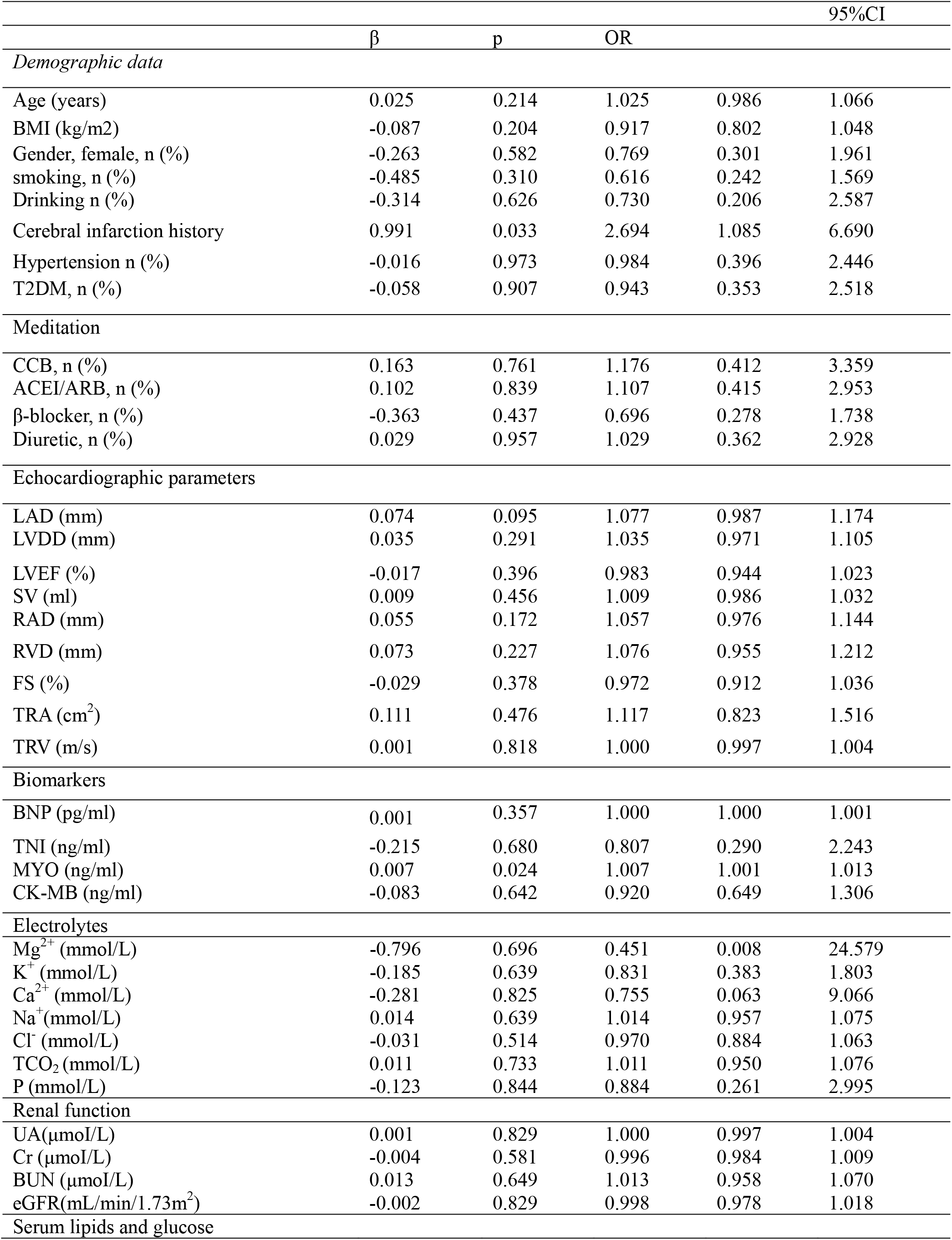
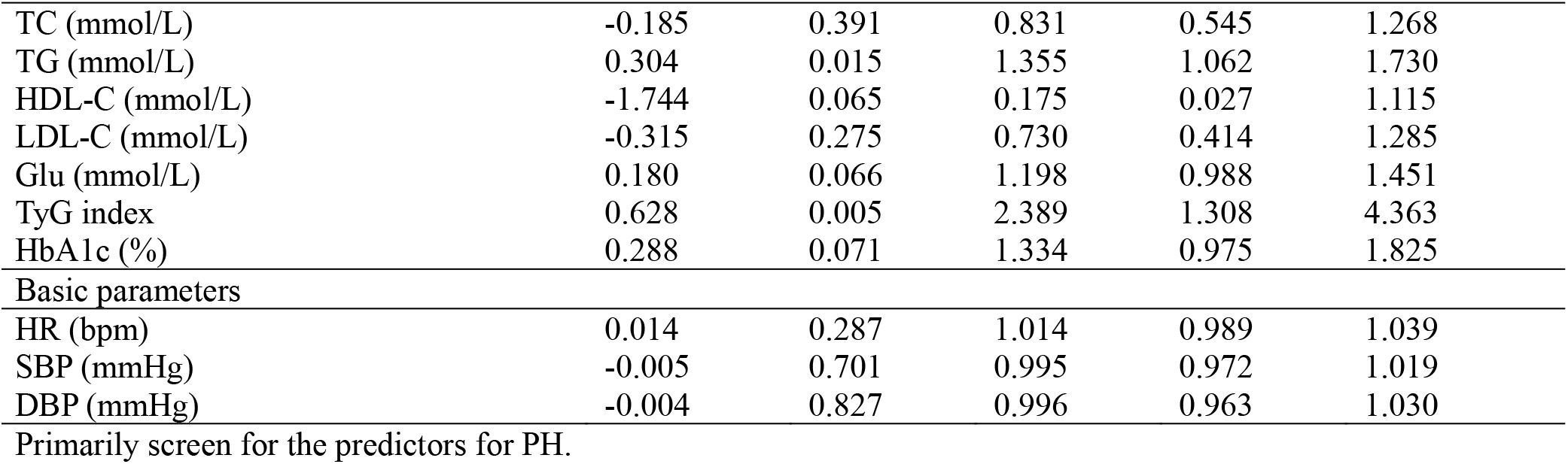
Univariate logistics regression for PH

| | $\beta$ | p | OR | 95%CI | |
| --- | --- | --- | --- | --- | --- |
| Demographic data |  |  |  |  |  |
| Age (years) | 0.025 | 0.214 | 1.025 | 0.986 | 1.066 |
| BMI (kg/m2) | -0.087 | 0.204 | 0.917 | 0.802 | 1.048 |
| Gender, female, n (%) | -0.263 | 0.582 | 0.769 | 0.301 | 1.961 |
| smoking, n (%) | -0.485 | 0.310 | 0.616 | 0.242 | 1.569 |
| Drinking n (%) | -0.314 | 0.626 | 0.730 | 0.206 | 2.587 |
| Cerebral infarction history | 0.991 | 0.033 | 2.694 | 1.085 | 6.690 |
| Hypertension n (%) | -0.016 | 0.973 | 0.984 | 0.396 | 2.446 |
| T2DM, n (%) | -0.058 | 0.907 | 0.943 | 0.353 | 2.518 |
| Meditation |  |  |  |  |  |
| CCB, n (%) | 0.163 | 0.761 | 1.176 | 0.412 | 3.359 |
| ACEI/ARB, n (%) | 0.102 | 0.839 | 1.107 | 0.415 | 2.953 |
| β-blocker, n (%) | -0.363 | 0.437 | 0.696 | 0.278 | 1.738 |
| Diuretic, n (%) | 0.029 | 0.957 | 1.029 | 0.362 | 2.928 |
| Echocardiographic parameters |  |  |  |  |  |
| LAD (mm) | 0.074 | 0.095 | 1.077 | 0.987 | 1.174 |
| LVDD (mm) | 0.035 | 0.291 | 1.035 | 0.971 | 1.105 |
| LVEF (%) | -0.017 | 0.396 | 0.983 | 0.944 | 1.023 |
| SV (ml) | 0.009 | 0.456 | 1.009 | 0.986 | 1.032 |
| RAD (mm) | 0.055 | 0.172 | 1.057 | 0.976 | 1.144 |
| RVD (mm) | 0.073 | 0.227 | 1.076 | 0.955 | 1.212 |
| FS (%) | -0.029 | 0.378 | 0.972 | 0.912 | 1.036 |
| TRA (cm<sup>2</sup>) | 0.111 | 0.476 | 1.117 | 0.823 | 1.516 |
| TRV (m/s) | 0.001 | 0.818 | 1.000 | 0.997 | 1.004 |
| Biomarkers |  |  |  |  |  |
| BNP (pg/ml) | 0.001 | 0.357 | 1.000 | 1.000 | 1.001 |
| TNI (ng/ml) | -0.215 | 0.680 | 0.807 | 0.290 | 2.243 |
| MYO (ng/ml) | 0.007 | 0.024 | 1.007 | 1.001 | 1.013 |
| CK-MB (ng/ml) | -0.083 | 0.642 | 0.920 | 0.649 | 1.306 |
| Electrolytes |  |  |  |  |  |
| Mg<sup>2+</sup> (mmol/L) | -0.796 | 0.696 | 0.451 | 0.008 | 24.579 |
| K<sup>+</sup> (mmol/L) | -0.185 | 0.639 | 0.831 | 0.383 | 1.803 |
| Ca<sup>2+</sup> (mmol/L) | -0.281 | 0.825 | 0.755 | 0.063 | 9.066 |
| Na<sup>+</sup> (mmol/L) | 0.014 | 0.639 | 1.014 | 0.957 | 1.075 |
| Cl<sup>-</sup> (mmol/L) | -0.031 | 0.514 | 0.970 | 0.884 | 1.063 |
| TCO<sub>2</sub> (mmol/L) | 0.011 | 0.733 | 1.011 | 0.950 | 1.076 |
| P (mmol/L) | -0.123 | 0.844 | 0.884 | 0.261 | 2.995 |
| Renal function |  |  |  |  |  |
| UA(μmol/L) | 0.001 | 0.829 | 1.000 | 0.997 | 1.004 |
| Cr (μmol/L) | -0.004 | 0.581 | 0.996 | 0.984 | 1.009 |
| BUN (μmol/L) | 0.013 | 0.649 | 1.013 | 0.958 | 1.070 |
| eGFR(mL/min/1.73m<sup>2</sup>) | -0.002 | 0.829 | 0.998 | 0.978 | 1.018 |
| Serum lipids and glucose |  |  |  |  |  |
| TC (mmol/L) | -0.185 | 0.391 | 0.831 | 0.545 | 1.268 |
| TG (mmol/L) | 0.304 | 0.015 | 1.355 | 1.062 | 1.730 |
| HDL-C (mmol/L) | -1.744 | 0.065 | 0.175 | 0.027 | 1.115 |
| LDL-C (mmol/L) | -0.315 | 0.275 | 0.730 | 0.414 | 1.285 |
| Glu (mmol/L) | 0.180 | 0.066 | 1.198 | 0.988 | 1.451 |
| TyG index | 0.628 | 0.005 | 2.389 | 1.308 | 4.363 |
| HbA1c (%) | 0.288 | 0.071 | 1.334 | 0.975 | 1.825 |
| Basic parameters |  |  |  |  |  |
| HR (bpm) | 0.014 | 0.287 | 1.014 | 0.989 | 1.039 |
| SBP (mmHg) | -0.005 | 0.701 | 0.995 | 0.972 | 1.019 |
| DBP (mmHg) | -0.004 | 0.827 | 0.996 | 0.963 | 1.030 |

Primarily screen for the predictors for PH.

Finally, the adjusted logistic regression indicated that cerebral infarction history (β = 0.990, OR: 2.690, 95% CI: 1.054-6.871, p = 0.039), MYO (β = 0.006, OR: 1.006, 95% CI: 1.000-1.012, p = 0.044), and TyG (β =0.636, OR: 1.889, 95% CI: 1.006-3.546, p = 0.048) were independent predictors of the onset of PH (table 6).

**Table 6.** Adjusted logistic regression for PH in CAD patients after PCI treatments

| | $\beta$ | p | OR | 95%CI | |
| --- | --- | --- | --- | --- | --- |
| Cerebral infarction history | 0.990 | 0.039 | 2.690 | 1.054 | 6.871 |
| TyG index | 0.636 | 0.048 | 1.889 | 1.006 | 3.546 |
| MYO | 0.006 | 0.044 | 1.006 | 1.000 | 1.012 |

Three of the above parameters were independent predictors of PH

## 4 Discussion

We performed a prospective cohort study within a median of 4.5 years’ follow-up. The primary endpoint focused on the onset of PH, which is frequently accompanied with various cardiovascular diseases. However, there have been few studies focusing on PH in CAD patients who have received PCI treatment. We analyzed the associations between the baselines of demographic data, renal functions, biomarkers for cardiac injury, serum lipids, and glucose. We have identified that the history of cerebral infarction, the TyG index baseline, and the MYO baseline were independent predictors for PH in CAD patients who received PCI treatment after 4.5 years.

### 4.1 PH in CAD patients after treatment with PCI

According to the previous guidelines and studies, PH refers to mean PAP (mPAP) >25mmHg at sea level measured by right heart catheterization (22). However, in the latest guidelines, the cutoff value of mPAP for PH has been updated to 20 mmHg, which may include more PH patients who were not diagnosed as PH in the past (22). Thus, a higher incidence of PH will be diagnosed in various diseases, and the early diagnosis and prevention of PH will prevent it developing into the advanced stage. The importance of PH in various cardiovascular diseases has been promoted. However, the diagnosis of PH evaluated by echocardiography has not yet been updated (26). Thus, in our present study, the cutoff of sPAP measured by echocardiography had not been revised and updated, and needs to be further studied in clinical practice. In future studies, we may investigate PH in accordance with the latest guidelines for the cut-off of sPAP evaluated by non-invasive echocardiography.

In our present study, the PH which accompanied CAD may be classified as PH due to left heart diseases (19,22,27). Left heart-related diseases are one of the most frequent causes of PH. However, the clinical prevalence data of left heart disease-related PH has not been uncovered (19,27). The incidence of PH has varied in different studies due to differing evaluation methods and criteria for PH diagnosis.

As discussed above, PH has been recognized as one of the most frequent complications of various cardiovascular diseases (28-31). However, in CAD patients, few studies have focused on PH complications. Incidence of PH was 8.91% among 247 CAD patients who received PCI treatment in our study. This incidence may be much higher if the cutoff is updated in the future (22). Though PH was not as significant as heart failure in CAD patients, it may affect right heart function or induce symptoms. PH is a common complication of CAD patients which has previously not been widely investigated and treated. Thus, PH should be given sufficient attention in clinical practice for CAD patient treatment and management.

### 4.2 Association of IR/TyG index and cardiovascular diseases

It has been indicated that IR is involved in the pathogenesis of various of diseases including atherosclerotic cardiovascular diseases (2). Diabetes is considered a primary risk factor of cardiovascular diseases, however, the association between diabetes and PH has not been confirmed (32). Though IR has been widely indicated to be a risk factor for a number of cardiovascular diseases including MACE and atherosclerosis (1,2,12,33), few studies have reported the roles of IR in PH, especially in CAD patients. The novel TyG index, a simple surrogate marker of IR, has been widely used as a marker/risk factor/predictor for a large number of cardiovascular diseases (5,34-36). A higher TyG index has been indicated to be correlated to hypertension (37). They found that the hypertension was significantly more likely in the patients with the highest level of TyG index than in the lowest level of TyG index patients (38). However, these results should be studied in future large-scale prospective cohort studies. In addition, further studies are needed to elucidate the potential pathophysiological mechanisms underlying the association between the TyG index and hypertension. Increasing evidence indicates that a higher TyG index or a higher level of TG and FBG levels increase atherosclerosis risk, including peripheral artery disease and arterial stiffness (39).

Triglycerides have been indicated to be independent risk factors for CVD. Uncontrolled diabetes and/or hyperlipidemia has been indicated to result to high level of TyG index, which is associated with symptomatic patients’ CVD (35).

### 4.3 TyG and PH in CAD patients after PCI treatment

Few researchers have focused on the complications of pulmonary vessels. Though PH has attracted an increasing amount of attention from clinical physicians and researchers, PH’s clinical characteristics and risk factors in CAD patients who have received PCI treatment have not been widely investigated (19,20,24,25,40). The roles of the TyG index in outcomes of patients who underwent PCI treatment have been partly reported (7,10,41,42). The associations between TyG index and poor prognosis in patients with acute ST-elevation myocardial infarction after PCI have also been indicated in previous studies(7,10). However, the potential role of the TyG index as a predictor of PH in CAD patients after PCI treatment has not been studied. Thus, we focused on the predictive role of baseline TyG for PH in CAD patients who have received PCI treatment, within a median of 4.5 years’ follow-up. In the present study, PH patients were indicated to be characterized with a higher level of TyG index, but this was not statistically significant. In the univariate regression, we found that the TyG index was a potential predictive factor of PH. Additionally, the adjusted regression indicated that the higher baseline TyG index was an independent predictor for the onset of PH in CAD patients who underwent PCI treatment. Though no evidence indicated that PH is caused by IR or abnormal metabolism of lipids or glucose, PH may be the results of IR in cardiovascular diseases (1,2). The patients in our study were CAD patients who may have also suffered from left heart failure, and the PH found may have been caused by this left heart failure.

Another possibility is that the IR or TyG index may be associated with PH via its roles in the heart or coronary arteries (43-45). Furthermore, it is also possible that IR has a detrimental effect on pulmonary vessels, increasing the remodeling of pulmonary vessels and PVR (44,45).

In clinical practice, a physician usually examines fasting glucose level to identify patients with high-risk, especially cardiovascular disease patients (11,46). The TyG index may also be valuable for prognostic cardiovascular disease risk stratifications, and may have a higher diagnostic significance than FBG and TG levels (11,46). However, PH has been indicated to be a progressively and dynamically progresses disorder. Thus, the TyG index as a prognostic marker can be easily biased by hyperlipidemia and diabetes.

### 4.4 Other predictors for PH in CAD patients

It is of great importance to identify the risk factors/predictors of PH as a prognosis of CAD patients who underwent PCI treatment for effective prevention. We have identified that beside the TyG index, the history of cerebral infarction and the baseline level of MYO were also independent predictors for PH in CAD patients. To the best of our knowledge, this is the first study to identify that the history of cerebral infarction was an independent risk factor for PH in CAD patients. This may be explained by the comorbidity of CVD and PH. It is worth noting that some of the patients included in our study were at risk of CAD because they had a previous history of CVD. Furthermore, baseline MYO was also indicated to be an independent risk factor for the onset of PH in CAD patients after PCI treatment. The above results suggest that myocardial injury may lead to chronic left heart failure, which is a common reason for PH.

## 5 Conclusion

PH is prevalent in CAD patients after PCI treatment. The baseline TyG index, cerebral infarction history, and MYO level were independent predictors for PH in CAD patients after PCI treatment.

## Data Availability

The data were available from the corresponding authors.

## 6 Conflict of Interest

We declare that the research was conducted without any commercial or financial relationships that could be construed as a potential conflict of interest.

## 7 Author Contributions

DQ and SZB participated in the design of the study. The manuscript and the statistical analyses were performed by LX, DQ and SZB. DQ reviewed and revised this manuscript critically for important intellectual content. Echocardiography examinations were performed by our technician RR. The sPAP-related assessments were recorded and analyzed by YX and LX. Clinical data and laboratory examinations data were performed and recorded by LX, LR and JL. Other laboratory data and patients’ follow-up were carried out by JC, YX and SF. The CAG operations were performed by LX, JC and DQ.

## 8 Funding

The present research is supported by the “Senior Medical Talents Program of Chongqing for Yong and Middle-aged” to Shi-Zhu Bian, “Miaopu” Young Scientists Program from Army Military Medical University (2019R046) and Basic Research Program from Army Military Medical University (2020D050).

## 9 Acknowledgments

The authors would like to thank all the individuals who participated in this study for their supports.

## 11. Abbreviations

AF: arial fibrillation
ACEI: angiotensin converting enzyme inhibitor
ARB: angiotensin receptor blocker
BNP: B-type natriuretic peptide
BUN: urea nitrogen concentration
CAD: coronary artery disease
CAG: coronary angiography
Ca2+: serum calcium concentration
CCB: calcium channel blockers
CI: Confidence interval
CK-MB: Creatine kinase-MB
Cl^−^: chloride ion
Cr: concentration of plasma creatinine
DBP: diastolic blood pressure
FBG: fasting blood glucose
FS: fractional shortening
HDL-C: high-density lipoprotein cholesterol
HR: heart rate
IR: insulin resistance
K+: serum potassium concentration
LAD: end-diastolic internal diameter of the left atria
LDL-C: low-density lipoprotein cholesterol
LVDD: end-diastolic internal diameter of left ventricle
LVEF: ejection fraction of left ventricle
LVPW: left ventricular posterior wall
MACE: major adverse cardiovascular events
MACCE: major adverse cardiac and cerebrovascular events
MYO: myoglobin
Na+: sodium concentration
OR: odds ratio
P: phosphate concpentration
PAP: pulmonary arterial pressure
PH: pulmonary hypertension
PCI: percutaneous coronary intervention
RAD: end-diastolic internal diameter of right atria
RVD: end-diastolic internal diameter of right ventricle
SBP: systolic blood pressure
STEMI: ST elevation myocardial infarction
T2DM: diabetes mellitus type 2
TG: triglyceride
TNI: Troponin
TR: tricuspid regurgitation
TRA: TR area
TRV: TR velocity
TyG index: triglyceride-glucose index

## Notes

### Competing Interest Statement

The authors have declared no competing interest.

### Funding Statement

This study was supported by the Senior Medical Talents Program of Chongqing for Yong and Middle-aged, the Excellent Scientist Pool Program of Army Medical University ("Miaopu Program 2019R046") and Basic Research Program from Army Military Medical University (2020D050).

### Author Declarations

Our study conforms to the Declaration of Helsinki. The study has been approved by our ethical committee of Xinqiao Hospital, Third Military Medical University (Army Military Medical University). The patients were familiarized with the procedure and contents of the study and provided written informed consent.

